# Systematic development of an mHealth app to prevent healthcare-associated infections by involving patients: Participatient

**DOI:** 10.1101/2021.02.28.21252122

**Authors:** R.G. Bentvelsen, R. van der Vaart, K.E. Veldkamp, N.H. Chavannes

**Affiliations:** Department of Clinical Microbiology, Leiden University Medical Center, Albinusdreef 2, 2333 ZA Leiden, the Netherlands; Microvida Laboratory for Microbiology, Amphia Hospital Breda, Molengracht 21, 4818 CK Breda, the Netherlands; Unit of Health, Medical and Neuropsychology, Faculty of Social and Behavioural Sciences, Leiden University, Wassenaarseweg 52, 2333 AK Leiden, the Netherlands; Department of Public Health and Primary Care, Leiden University Medical Center, Albinusdreef 2, 2333 ZA Leiden, the Netherlands

**Keywords:** mHealth, Development, Patient engagement, Infection prevention and control, Catheter-associated urinary tract infections

## Abstract

**Introduction:** In hospital care, urinary catheters are frequently used, causing a substantial risk for catheter-associated urinary tract infections (CAUTI). Patient awareness and evaluation of appropriateness of their catheter through mHealth could decrease these healthcare-associated infections. However, patient engagement via mHealth in infection prevention is still limited. Therefore, we describe the systematic development and usability evaluation of the mHealth intervention Participatient, to prevent CAUTI, aiming for optimal adoption of the app in the clinical setting.

**Method:** The CeHRes roadmap was used as development guideline, operationalizing phases for (1) contextual inquiry (observations and interviews), (2) value specification (interviews with probing) and (3) design in multiple steps and in co-creation with end-users. During phases 1 and 2, semi-structured interviews were conducted with fifteen patients and three nurses. The design phase was combined with the minimum viable product development strategy, with a focus on early cyclic steps of prototyping.

**Results:** In phase 1, patients acknowledged the risks of catheter use. Patients in phase 2 valued endorsement of a mHealth application by healthcare workers and reported to own a smartphone. Both patients and nurses recognized the need for useful modules in the app besides catheter care. Based on the needs and values as found in phase 2, the Participation app was developed. Based on usability tests in phase 3, content, text size, plain language, and navigation structures were further amended, and images were added.

**Conclusion:** This study provides real-world insight in the developmental strategy for mHealth interventions by involving both patients and care providers. Development of an app using thorough needs-assessment provided understanding for its content and design. By developing an app providing patients with reliable information and daily checklists, we aim to provide a tailored tool for communication and awareness on catheter use for the whole ward, and a potential blueprint for mHealth development.

## 1. Introduction

Within the hospital setting, healthcare-associated infections (HAI) are a high-priority problem as they are associated with an increased mortality, more adverse events, and increased length of stay in the hospital or long-term care facility.^1, 2^ The most prevalent HAI is the catheter-associated urinary tract infection (CAUTI), which is best prevented by reducing the use of urinary catheters.^1-4^ Previous studies have shown that 20 - 50% of urinary catheters are placed with an inappropriate indication^5^ or are not timely removed.^1, 4^ Reduction of CAUTI has become a priority by the European Centre for Disease Prevention and Control (ECDC) and national institutes like the Dutch National Institute for Public Health and the Environment (RIVM).

The current approach in reducing catheter use, and thus CAUTI, is creating awareness among physicians and nursing staff by implementing guidelines and surveillance with feedback.^2, 6^ However, this approach is considered time consuming, financially expensive, with declining compliance over time, and insufficient reduction of inappropriate catheter placement.^7, 8^ To overcome these barriers, a relevant addition to reach catheter reduction would be to involve the patients in the decision making process regarding the catheter removal.^9^

With increasing use of mobile health applications (apps), new tools become available for patient engagement and self-management. Such mHealth technologies have shown potential to improve safety and quality of care. For example, by providing patients with tailored information on their medical condition or as a means for communication on their health.^10, 11^ A review of the most used app stores in 2015 reported seventeen apps available regarding HAI prevention, of which a mere two apps were also aimed at patients.^13^ One of them, called “Infection Prevention” is an informative app, listing precautions associated with preventing numerous infections. The other is the “Handwash counter”, which lets patients and providers track handwashing instances to promote infection prevention and better hand hygiene. These apps are pioneering in the field of infection prevention, though the number of downloads and thus the use of these apps is still limited. Furthermore, scientific evidence for the effectiveness of these apps is lacking.

The main obstacles in mHealth adoption are limited usefulness to the patient, poor functionality and usability, and lack of integration in clinical routines.^13, 14^ Usability defines the extent to which a product, system or service can be used by specific users to achieve specific goals with effectiveness, efficiency, and satisfaction in a specific context of use (definition from ISO standard 9241.18). Reducing these obstacles could lead to higher adoption rates in clinical settings, more effectiveness and efficiency and higher satisfaction of users.^3, 14^ To reach optimal adoption, a structured approach as described in the CeHRes roadmap could be used, including focus on contextual inquiry, value specification, and design and prototyping.^15^ Additionally, a widely used method for early testing is described in the Minimum Viable Product (MVP) strategy. This provides unbiassed insight with a short build-measure-learn feedback loop, creating a process in which end-users are optimally involved in technology development.^16^

In the current paper, the development of an infection prevention mHealth intervention is described, aiming to involve patients in the decision-making process around their urinary catheter. The development follows the CeHRes roadmap and the MVP strategy. The intention in using this approach is to learn how to build a sustainable intervention and promote patient-provider communication in infection prevention. Additionally, we aim to set a blueprint for mHealth development, with valuable lessons learned, which could be translated into a broad clinical context.

## 2. Method

### Study design

In the developmental process, two nursing wards with high urinary catheter use and adult patients were included, at the Leiden University Medical Center, the Netherlands. One ward for general surgery and orthopedics, the second ward for solid organ transplant patients. From the CeHRes Roadmap the phases (1) ‘contextual inquiry’ (CI), (2) ‘value specification’ (VS) and (3) ‘design’ were deployed, between April and June 2017 (Figure 1). In the design phase the CeHRes roadmap was combined with the MVP strategy. CI and VS were operationalized using interviews with both nurses and patients. Nursing staff were asked to indicate which patients could be approached for inclusion, excluding patients impaired through illness either physically or mentally. During all patient interactions (interviews, observations, and interactions with the app), patients were asked for informed consent, at the patient’s bedside or in a separate consultation room on the ward.

**Figure 1.**
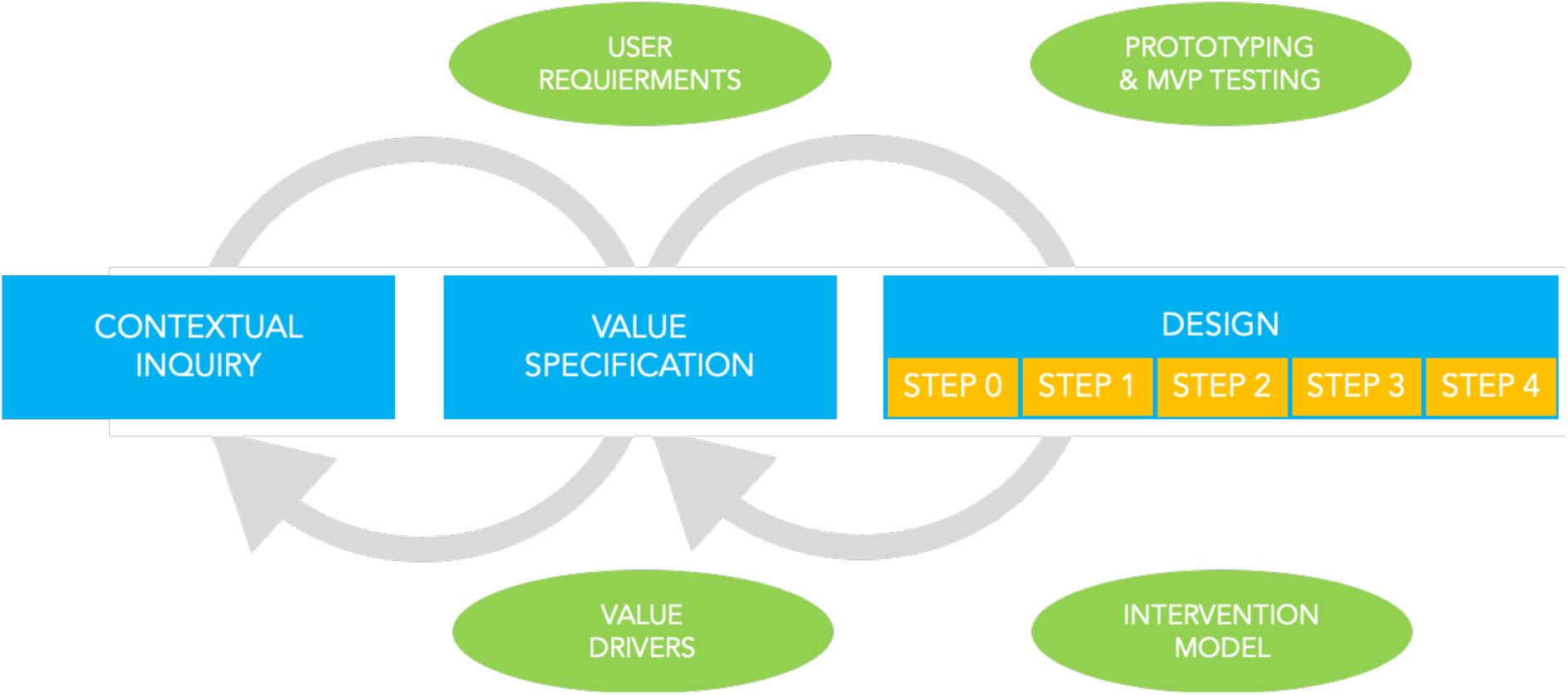
The Participatient development roadmap. The Participatient development roadmap is based on the CeHRes roadmap and the MVP strategy. A set of essential requirements gathered in the contextual inquiry and value specification phases was used in the design phase, with MVP testing in pre-clinical (step 0) and end-user testing steps. The operationalization and summative evaluation phases are discussed in a follow-up study.

The project was coordinated by a multidisciplinary project management team with researchers and healthcare professionals from different fields, namely infection prevention (RB, KV), psychology (RV), and eHealth development (NC). In addition, there was close collaboration with designers and engineers.

### 2.1 Contextual Inquiry

The first phase of the CeHRes roadmap comprises a contextual inquiry (CI). To understand the context of the hospital care on different wards and to understand the needs for a patient centered CAUTI prevention intervention, field observations of habits and rituals around hospital admission, and catheter-use and removal were conducted. Subsequently, the observational CI was supplemented with semi-structured interviews with patients and nurses (Appendix A). Questions regarded (1) demographics, being age group, ward of admission, and sex; and (2) potential (perceived) barriers for app use among patients, such as motor skill or sight impairments. To check for the present level of eHealth use (3), semi-quantitative questions were posed on (A) smartphone use, (B) health-related app use, and (C) internet queries for health-related questions. For the catheter-infection prevention aspects of this project, several questions were posed on (4) urinary catheter care experiences, namely (A) the perceived quality of the urinary catheter care, (B) the level of patient-provider communication concerning the indication and need of a catheter, and (C) the awareness on the risk for catheter-associated urinary tract infection.

### 2.2 Value specification

The second phase according to the CeHRes roadmap concerns the value specification. The development team started out defining essential values for the app that were to be taken into account. Within the same interviews as described for the contextual inquiry interviewees were asked about their specific values for an mHealth intervention (Appendix A). The goal was to create a rich picture of the expected triadic interaction between patient, professional and intervention, in order to meet the needs of patients and staff. With exploratory questions patients and nursing staff were asked to elaborate on their view on the needs regarding the app. Questions regarded (1) reasons to use a hospital admission app, and (2) interviewees views on them using it, and (3) how to create optimal involvement of patients in their healthcare process during admission, (4) especially in urinary catheter care, (e.g. by using motivational messages to discuss the indication, reminders to perform the catheter check, and scientific proof on indications); and (5) what requirements a hospital admission app should meet for them to use it, e.g. practical information on the admission.

### 2.3 Design and Prototyping

In the third phase of the CeHRes roadmap, the design phase, medical students and nurses primarily helped as volunteers to test and correct basic functionality of the app and to check for complex medical phrases. Subsequently, the app was tested with patients. Prototyping started early in the development of the app, using the MVP strategy. The MVP is a first version of a new product which allows a team to collect maximum feedback of end-users in the most efficient manner. By testing the app multiple times and from a very early stage, the feedback loop is shortened and flaws or omissions are detected early on to re-iterate towards the optimal design (Appendix A).^16^ Typically, four short cyclic steps are taken in the design process, which translate functional requirements from the CI and VS phases into a minimum set of specifications built into two iterations (version A and B) of a product. The same tester (patient or nurse) evaluates both versions for comparison. Testers were asked to use their own mobile device, as issues can remain undetected due to false assumptions about technology available to patients.^12^ Each of the four design steps involved concurrent probing and subjects were invited to ‘think aloud’ during evaluation of the app.^17^ During testing, the researchers used concurrent probing on the design regarding: (1) readability of the text; (2) usefulness of the graphics; (3) ability to start and use the app without help; (4) explanation of medical terms; (5) purpose of the app according to the user; and (6) an overall rating of the app. After each step, the app was adjusted based on the evaluations of the A and B versions, resulting in new features for the next evaluation step, ending with the choice for the essential and most popular features, which were developed in full.

In the second and third design step, after testing participants filled out an app-specific questionnaire based on the MARS, to assess the perceived impact of the app on the user’s attitudes, awareness, knowledge, intentions to change, help seeking behavior as well as the likelihood of actual change in the target behavior.^18^ In the third step of app testing, usability was additionally assessed using the System Usability Scale (SUS).^19^ This industry standard is a subjective evaluation in 10 questions with a five-point Likert scale. The SUS results in a score ranging from 0-100, with a score above 67 qualifying as above average.^20^

## 3. Results

### Demographics

In total, 28 people were involved in CI, VS, and the design phase, of which 22 were patients. These patients were all above 18 years old, for privacy reasons age was listed per age group. Eight (36.4%) patients were in the youngest age group (18-44 years old), followed by six (27.3%) aged between 45 and 59, and seven (31.8%) of 60–74. One (4.5%) patient was older than 74 years. Of all patients, 31.6% (7) were male and 68.4% (15) female. Ten of the patients were admitted on the transplantation ward and twelve on the combined ward for orthopedics and general surgery. Of the 22 patients, 15 participated in both the interviews for CI and VS phases and the early prototypes. Three nurses participated in the interviews for CI, VS and early prototyping testing.

### 3.1 Contextual Inquiry

The interviewed patients and nurses indicated that they and (other) patients typically own and carry a smartphone during their hospital stay. Visual impairments as potential limitation for app use were reported by four out of fifteen (27%) patients, of which half were related to the hospital stay (e.g. surgery or anesthesia). Motor skill impairment was noted by 20% of patients (three out of fifteen). Impairments to smartphone use during admission were mostly mild and temporary, for example somnolence due to the surgery or anesthesia. All of the respondents had internet access on their device. Five out of the thirteen respondents who answered this question (38%) had an iPhone versus eight (62%) Android users. Ten out of fourteen (71%) of respondents searched online for health-related questions, and 43% (6 out of 14) used apps related to health.

The field observations and interviews with nurses on urinary catheter use, communication on the indication, and the risks for CAUTI exposed three key moments for intervention. The first is conscious catheter indication before placement. Second is periodic (written) reflection on the reason for the catheter during placement, which, according to the nurses was often not a priority in routine care by nurses or physicians. Third, the barriers for removal of the catheter are more direct than the indirect potential risk. For example, the ease of care with a catheter in place versus the indirect and protracted risk of CAUTI. Based on these three key factors, it was decided that a checklist to check on the appropriateness of a patient’s catheter – the Catheter Check – in the app could promote awareness on the reason for the catheter and the potential risk of CAUTI among patients and staff. The questions in this Catheter Check could and must be based on the current practice guidelines for catheter indications.

### 3.2 Value Specification

The pre-set values (0) for the Catheter Check in the app, as defined by the development team stakeholders were: (1) adherence with current guidelines on urinary catheter use, (2) suitability for all age groups and skill levels, (3) support regarding self-management – patients should be able to find, install, and use the app on their own, and (4) enabling patients to participate in the communication and decision making regarding their urinary catheter.

Nurses and patients indicated the importance of the correct moment of introduction of the app. Patients are often limited by their disease or treatment at the first day of hospital admission, so an introduction of the app on day two or three would be better. “I would use the app if it was free and reliable”, one respondent answered.

Patients indicated that for optimal involvement, if a staff member would recommend the app, they would be more likely to use it. By adhering to the evidence-based guidelines, caregivers are more likely to promote the app. “A clear call to action after the catheter checklist conclusion would help”, a patient reported who had experienced a stressful time with a dermatological app without advice after the presumed diagnosis. An older interviewee pointed out the value of involvement: “It is important to … instruct users to discuss findings with their doctor or nurse”, and “the app could help to explain the reason for the catheter”.

Patients asked for plain language with a simple login procedure, suitable for self-management in all patients. The interviews showed that the most frequently asked questions by patients regarded practical information on the hospital admission. This would be an important requirement for them to use a hospital app. This information should include information on the room; visiting hours, route and phone numbers of the hospital and ward; and the route to the pharmacy. Additionally, we asked physicians of various specialties for input, and the department of anesthesiology asked for collaboration to create a digital pain score monitoring tool.

### 3.3 Design and Prototyping

From the findings in the contextual inquiry and value specification phases, an app prototype was developed containing a catheter check, information on the hospital and ward, medical information, and a pain score monitoring tool (Appendix C). The catheter check was developed regarding the purpose of preventing CAUTI through reducing inappropriate urinary catheter use. This makes the current guidelines for appropriate urinary catheter use ^21^ accessible for patients. The catheter check in the app contains a brief introduction on catheters and risks, after which the app displays an interactive checklist of 8 questions with simple answer options, to monitor the catheter indication and appropriateness. Based on these eight questions, a valuation of the appropriateness of the catheter is provided (appropriate, inappropriate, or unknown). If the catheter placement reason is scored to be inappropriate or unknown, the user receives messages of encouragement to discuss this with their nurse or physician in a conversation during ward rounds or ad hoc. The app was made freely available with instructions for download via a leaflet, which patients received with recommendation from their nurse at admission. No privacy sensitive questions were asked when entering the app, the user simply needed a hospital ward code, as provided on the leaflet.

After pre-clinical testing with medical students and nurses, patients tested two versions of the app in one of the four design steps. The design testing steps led to several changes in the intervention (Appendix 3). Text size was increased, and syntax and grammar used were simplified. Illustrations were added to help clarify the text. Downloading and starting the app required assistance at times, so the instruction leaflet was improved. Medical vocabulary was explained more carefully after a comment from a patient: “Straight off first mentioning the risks is kind of intense. It might be better to first mention that you do not always have a choice, and what the consequences of that could be. That is just a little less heavy”. Other patients mentioned they would ask their physician for clarification of medical terms if needed. The goal of the app was clear for all testing nurses and patients. The app improved per version, as the testers rated version 2a with an average 7.0 (n=6), and 2b a mere 6.0 (n=4). 3a was rated the best version with an average of 7.6 out of ten (n=6), (see Appendix for details). The app-specific questionnaire indicated the perceived impact of the app versions increased from an average of 3.0 out of 5 in version 2a, through 3.9 in 2b and 4.2 in version 3.From the SUS scores we learnt version 3ascored good with 72 out of 100 points where 2b scored a marginal 68 points. The final app 3b was built on the designs from version 3a and no usability issues were reported in the last tests (Appendix B).

## 4. Discussion

In this paper, the development of an infection prevention mHealth application is described, by involving patients in the decision-making process around their urinary catheter with a checklist on the indication. The development follows a theoretically driven approach based on the CeHRes roadmap^15^ combined with early cyclic testing as described in the MVP strategy.^16^ This resulted in the development of a patient-centered CAUTI prevention app, called Participatient.

In the hospital setting, urinary catheter care is not a priority, according to the interviewed nurses, although they do acknowledge the risks for CAUTI. Literature shows good examples that patient involvement can play a major role in prioritizing the limitation of risks, which has been successful in hand hygiene care and in prevention of post-discharge surgical site infections.^22, 23^ In a thorough review by McGuckin et al. on patient empowerment and hand hygiene it is shown that, at least in principle, patients are willing to be empowered. This review concludes that the actual performance of patient empowerment can be increased by providing patients with explicit permission and encouragement to be involved in the matter, by hospital staff.^24^ Up until now, in urinary catheter care patients are not included in such a manner yet. Our study shows that patients acknowledge the risks of infections and the benefits of involvement and that an app to support this involvement shows large potential. This indicates a basis for patient participation, however adoption of the app in clinical practice is subject for further research.

A fraction of the pioneering apps on (patient centered) infection prevention and hand hygiene make a great case to invoke the potential of patient participation, though the effects are not published in literature.^13^ The lack of eHealth implementation could be improved with a systematic development and focus on functionality, usefulness for the patient, usability and integration in clinical routine.^15^ The phases of contextual inquiry and value specification showed that an mHealth intervention to enhance patient engagement in CAUTI prevention should include a checklist for catheter appropriateness endorsed by medical staff in plain language, complemented with additional relevant information on their hospital stay. Moreover, the design phase showed that clear instructions for download and use, a large text font and illustrations are essential in order to make the app as usable as possible.

Limitations of this study could include the sample size and confined group of patients. However, the sample of five participants per round is described by Nielsen as sufficient for technological development and usability testing.^25^ The wards were chosen because of high catheter use and motivation to join the project. However, a specific patient population like this could limit the implications in different settings. Also, we did not register factors affecting smartphone use as educational level and (eHealth) literacy. However, in consultation with nurses we aimed to test in a representative population of the wards for this qualitative study. Moreover, patients and professionals involved in interviews are likely positively biased for innovations. This is nevertheless the group that would adopt an app first and set an example. We aimed to design the app as accessible as possible. Though some people will not be willing or able to use the app themselves. However, app use by their family, friends, care providers, and other patients on the ward will raise awareness regarding CAUTI for the benefit of the ward as a whole.

## Conclusion

This study provides real-world insight in the developmental strategy for mHealth interventions by involving both patients and care providers. Development of an mHealth application using thorough needs-assessment provided essential understanding for its content and design. Participatient has the potential to improve awareness on appropriate indwelling catheter placement per ward as a whole. By developing an app providing patients with reliable information and daily checklists, we aim to provide a tailored tool for communication and awareness on catheter use, and a potential blueprint for mHealth development projects. The results of this study could be used to further improve the adoption of mHealth in-hospital apps. In future studies, we plan to implement and test the effect on reducing hospital infections like CAUTI by involving hospitalized patients.

### Practice implications

- Safety and quality of care could be improved by patient engagement and increased communication on infection prevention. Inappropriate catheter use is a feasible test case for patient engagement in HAI infection prevention to reduce catheter-associated urinary tract infections.
- The implementation of mHealth in the hospital setting is challenging because of the patient population and their impairments. Contextual inquiry, value specification and testing the design with patients and nurses on wards is essential for the adoption and successful implementation of eHealth.
- For future research it is relevant to test feasibility of the Participatient concept and implementation of this intervention on hospital wards. The low amount of mHealth experience among smartphone users confirms that special attention needs to be payed to addressing the benefits from using mHealth in communication towards patients.

## Data Availability

Anonymized crude data is present at the department of Medical Microbiology, Leiden University Medical Center, The Netherlands and will be saved.

## Publication requirements

### Appendices

– APPENDIX A – Interviews & Prototyping
– APPENDIX B – Screenshots of the functional app
– APPENDIX C – Description of prototyping process with issues and actions taken

## Abbreviations

CAUTI: Catheter-associated urinary tract infection;
CI: Contextual inquiry
ECDC: European Centre for Disease Prevention and Control
HAI: Healthcare-associated infections
mHealth: Mobile electronic healthcare
MVP: Minimum Viable Product
RIVM: Dutch National Institute for Public Health and the Environment
VS: Value specification

## Authors’ contributions

RB, RV, KV, and NC made substantial contributions to the concept and design of the study, and the acquisition of data. NC contributed in the design of the study and facilitated the logistics and means to develop the app. RB acted as interviewer of patients and healthcare workers. RB subsequently analyzed the data and interpreted the data regarding the app and was a major contributor to the manuscript. RV, KV, and NC revised the manuscript critically for important intellectual content. All authors read and approved the final manuscript.

## Consent for publication

Patient and health care worker consent was obtained for this study, before interviews, observations or interactions with the app.

## Availability of data and materials

The datasets used and analyzed during the current study are available from the corresponding author on reasonable request.

## Ethics approval and consent to participate

The study has been waived from requiring ethics approval by the Medical Research Ethics Committee [MREC] of Leiden University Medical Center. I confirm all personal identifiers have been removed so the persons described are not identifiable and cannot be identified through the details of the story.

## Acknowledgements

We thank the patients and healthcare workers of the nursing wards of the LUMC, the Dutch *Hacking Health* (hacking-health.org/) hackathon, and the following researchers who have contributed to the project: Hélène Trommelen for logistics, interviews and designs, Marguerite Bruijning for advice on infection prevention implementations. Chris Martini for the Pain score module, and Lauwerens Metz and Innovattic B.V., who contributed to discussions on design, usability and implementation.

## Appendices

### APPENDIX A – Interviews & Prototyping

#### Interview questions used

##### Contextual Inquiry

1. Demographics of potential end-users: Nursing ward, Age group, Sex.
2. Impairments, permanent or temporary (e.g. related to admission): visual, motor skills.
3. eHealth skills evaluated through semi-quantitative frequency of internet use, smartphone use, mHealth use.
4. Urinary catheter use, experience, and knowledge on risks of CAUTI.

##### Value specification

1. What would you like in a hospital admission app?
2. What are reasons to use a hospital admission app?
3. How could an app help promote shared decision-making during admission?
4. Would you use a shared decision-making app on urinary catheters during admission?
5. What requirements should a hospital admission app meet?
6. How important is data safety and privacy for you, would this be a reason not to use the app?

##### Design and usability

1. Is the text readable in your opinion?
2. Do you like the graphical interface?
3. What is your opinion on the data entry method?
4. Do you think you would need help using the app (for the first time)?
5. Are the medical terms clear or explained enough to you?
6. What is the goal of the app in your words?
7. Would you rate the app from 1 till 10?

Behavior of subject during prototyping is transcribed when deviating from expected. For example, not clicking the designated word or button. Also, exclamations from the thinking aloud were noted.

**Table A.1.**
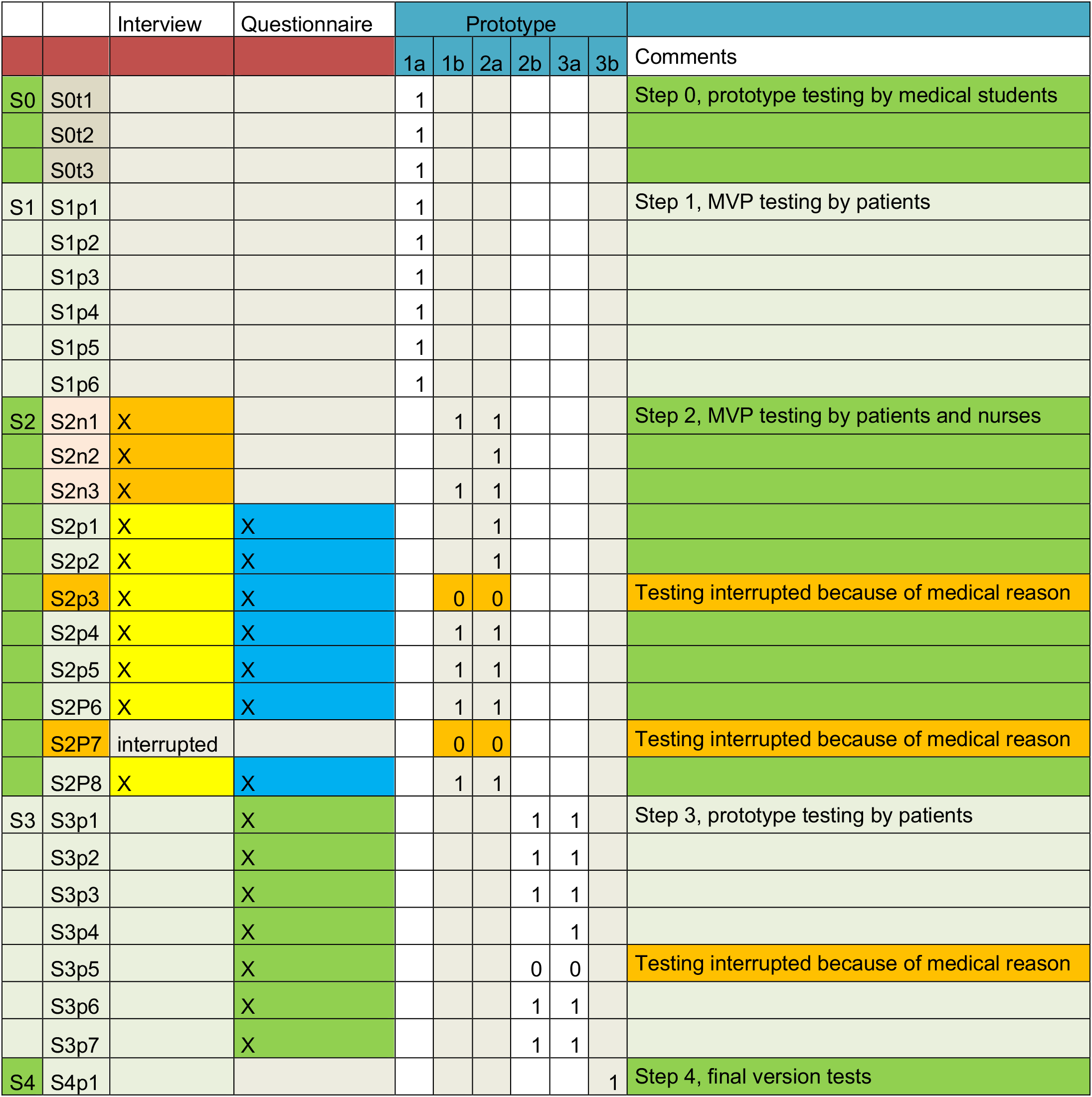
Interview and prototyping overview. In the various steps (S) of prototyping (S0 till S4) the prototypes (1a till 3b) were tested by a total of 3 medical student testers (t); 3 ward nurses (n); and 22 admitted patients (p) of which 3 prototype test sessions were excluded due to medical interruptions. The interruptions were due to ward rounds of physicians or medical check-ups. Also interviews in step 2 with the 3 nurses and 8 patients of which 1 patient interview was excluded from analysis due to interruption.

|  |  | Interview | Questionnaire | Prototype |  |  |  |  |  |  |
| --- | --- | --- | --- | --- | --- | --- | --- | --- | --- | --- |
|  |  |  |  | 1a | 1b | 2a | 2b | 3a | 3b | Comments |
| S0 | S0t1 |  |  | 1 |  |  |  |  |  | Step 0, prototype testing by medical students |
|  | S0t2 |  |  | 1 |  |  |  |  |  |  |
|  | S0t3 |  |  | 1 |  |  |  |  |  |  |
| S1 | S1p1 |  |  | 1 |  |  |  |  |  | Step 1, MVP testing by patients |
|  | S1p2 |  |  | 1 |  |  |  |  |  |  |
|  | S1p3 |  |  | 1 |  |  |  |  |  |  |
|  | S1p4 |  |  | 1 |  |  |  |  |  |  |
|  | S1p5 |  |  | 1 |  |  |  |  |  |  |
|  | S1p6 |  |  | 1 |  |  |  |  |  |  |
| S2 | S2n1 | X |  |  | 1 | 1 |  |  |  | Step 2, MVP testing by patients and nurses |
|  | S2n2 | X |  |  |  | 1 |  |  |  |  |
|  | S2n3 | X |  |  | 1 | 1 |  |  |  |  |
|  | S2p1 | X | X |  |  | 1 |  |  |  |  |
|  | S2p2 | X | X |  |  | 1 |  |  |  |  |
|  | S2p3 | X | X |  | 0 | 0 |  |  |  | Testing interrupted because of medical reason |
|  | S2p4 | X | X |  | 1 | 1 |  |  |  |  |
|  | S2p5 | X | X |  | 1 | 1 |  |  |  |  |
|  | S2P6 | X | X |  | 1 | 1 |  |  |  |  |
|  | S2P7 | interrupted |  |  | 0 | 0 |  |  |  | Testing interrupted because of medical reason |
|  | S2P8 | X | X |  | 1 | 1 |  |  |  |  |
| S3 | S3p1 |  | X |  |  |  | 1 | 1 |  | Step 3, prototype testing by patients |
|  | S3p2 |  | X |  |  |  | 1 | 1 |  |  |
|  | S3p3 |  | X |  |  |  | 1 | 1 |  |  |
|  | S3p4 |  | X |  |  |  |  | 1 |  |  |
|  | S3p5 |  | X |  |  |  | 0 | 0 |  | Testing interrupted because of medical reason |
|  | S3p6 |  | X |  |  |  | 1 | 1 |  |  |
|  | S3p7 |  | X |  |  |  | 1 | 1 |  |  |
| S4 | S4p1 |  |  |  |  |  |  |  | 1 | Step 4, final version tests |

### APPENDIX B – Screenshots of the functional app

**Figure B.1:**
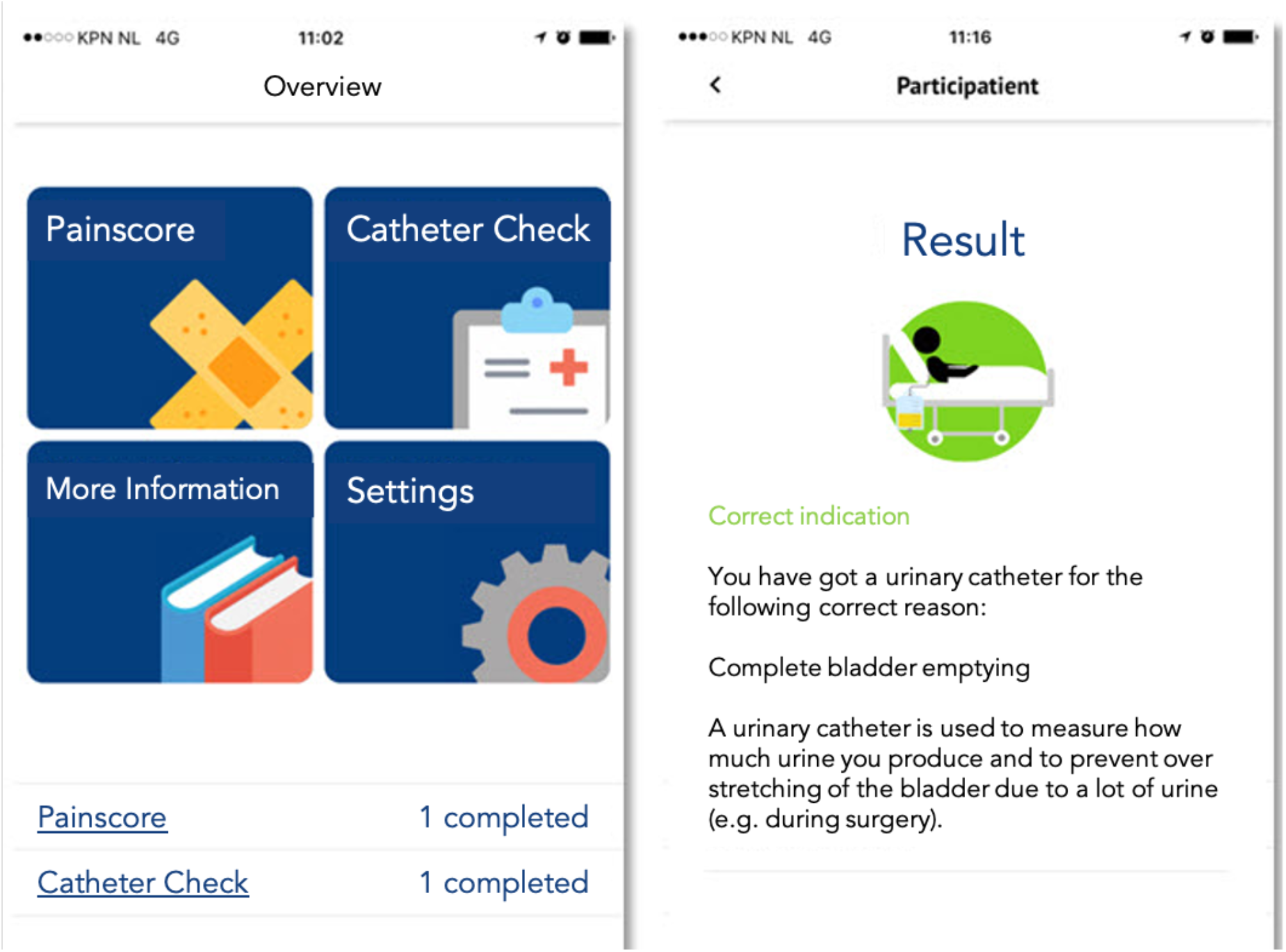
Screenshots of the Participatient app version 3 menu (left) and result screenshot (right).

**Figure B.2:**
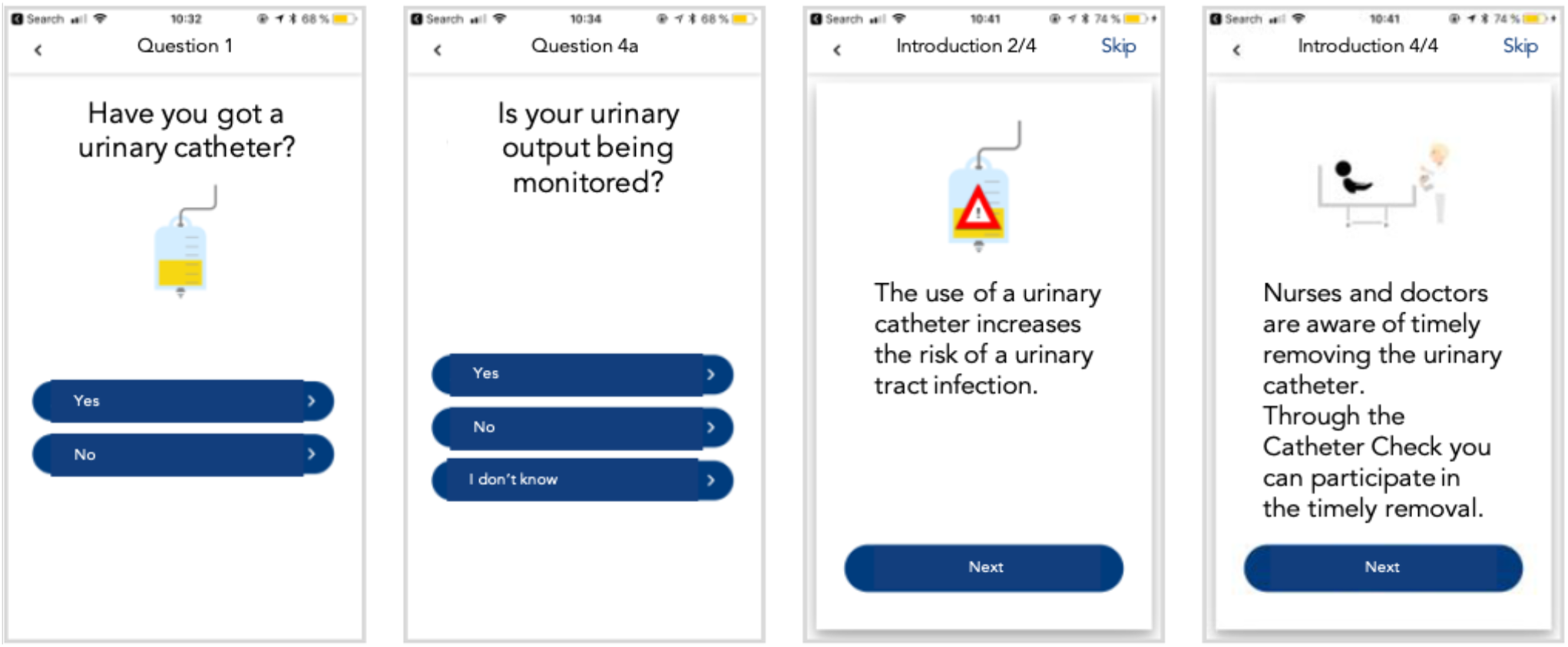
Screenshots of the Participatient app and Catheter Check version 3. From left to right: (a) Question 1 ‘Have you got a urinary catheter?’; (b) Question 4A ‘Is your urinary output being monitored?’; (c) Introduction text: ‘The use of a urinary catheter increases the risk of a urinary tract infection.’; (d) Introduction text: ‘Nurses and doctors are aware of the timely removal of the urinary catheter. Through the *Catheter Check* you can participate in the timely removal.’

### APPENDIX C – Description of prototyping process with issues and actions taken

The app versions tested, with focus on the Catheter Check part of the Participatient hospital admission app, with reported issues and actions taken are summarized in table C.11.

**Table C.1.**
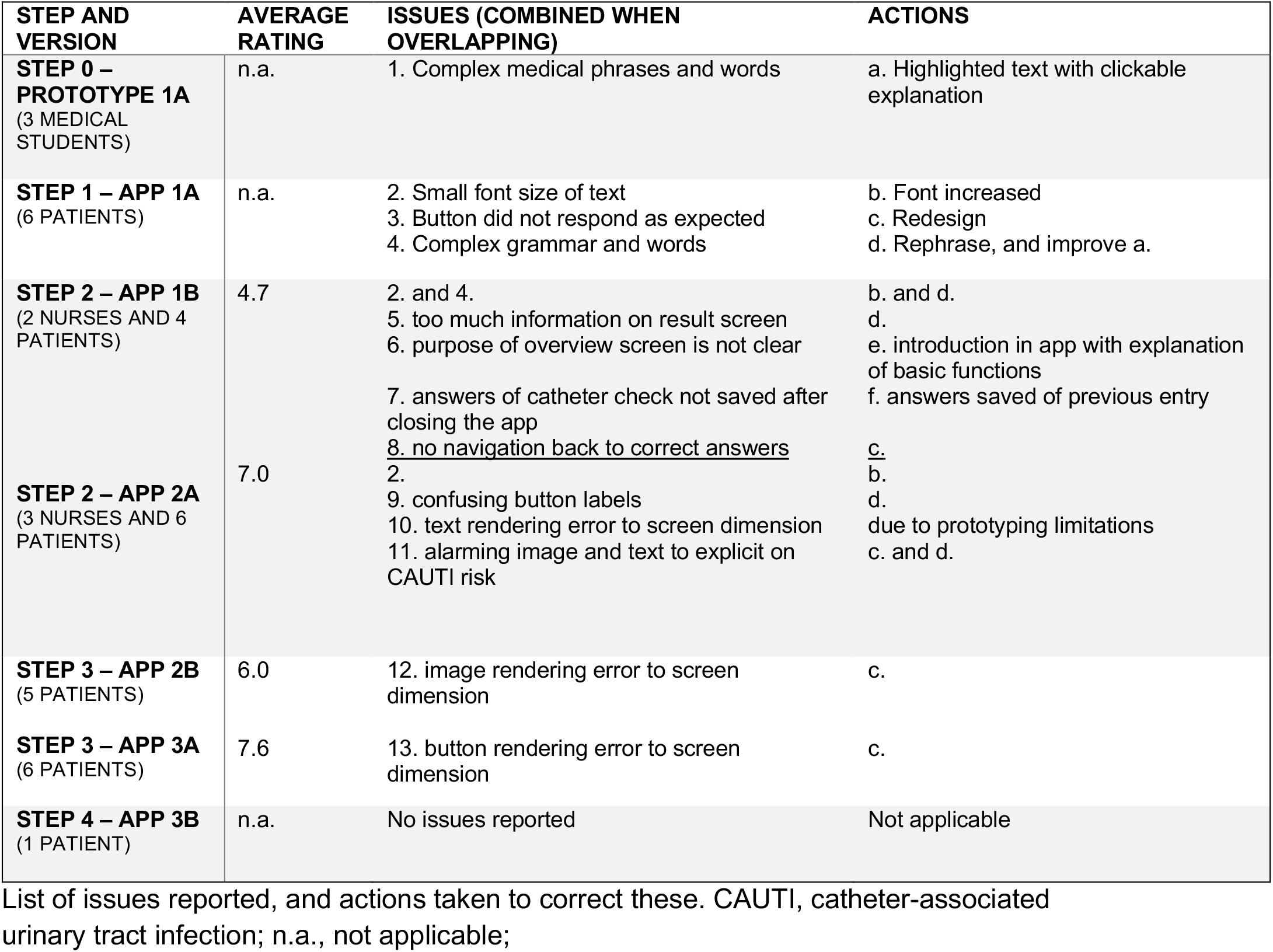
Qualitative description of prototyping process with issues and actions taken.

## References

1. Suetens C, Hopkins S, Kolman J, Diaz Högberg L. Point prevalence survey of healthcareassociated infections and antimicrobial use in European acute care hospitals. Stockholm: ECDC; 2013

2. Laan BJ, Spijkerman IJ, Godfried MH, Pasmooij BC, Maaskant JM, Borgert MJ, et al. De-implementation strategy to Reduce the Inappropriate use of urinary and intravenous CATheters: study protocol for the RICAT-study. BMC Infect Dis. 2017;17:53, doi.org/10.1186/s12879-016-2154-2.

3. Middleton B, Bloomrosen M, Dente MA, Hashmat B, Koppel R, Overhage JM, et al. Enhancing patient safety and quality of care by improving the usability of electronic health record systems: recommendations from AMIA. J Am Med Inform Assoc. 2013;20:e2–8, doi.org/10.1136/amiajnl-2012-001458.

4. Lorig KR, Holman H. Self-management education: history, definition, outcomes, and mechanisms. Ann Behav Med. 2003;26:1–7, doi.org/10.1207/S15324796ABM2601_01.

5. Werkgroep Infectiepreventie. Preventie van infecties als gevolg van blaaskatheterisatie via de urethra [Prevention of infactions caused by urethra catheterization]. Leiden: Leids Universitair Medisch Centrum; 2010

6. Van den Broek PJ, Wille JC, van Benthem BH, Perenboom RJ, van den Akker-van Marle ME, Niel-Weise BS. Urethral catheters: can we reduce use? BMC Urol. 2011;11:10, doi.org/10.1186/1471-2490-11-10.

7. Meddings J, Saint S. Disrupting the life cycle of the urinary catheter. Clin Infect Dis. 2011;52:1291–3, doi.org/10.1093/cid/cir195.

8. Harbarth S, Sax H, Gastmeier P. The preventable proportion of nosocomial infections: an overview of published reports. J Hosp Infect. 2003;54:258-66; quiz 321.

9. Marschang S, Bernardo G. Prevention and control of healthcare-associated infection in Europe: a review of patients’ perspectives and existing differences. J Hosp Infect. 2015;89:357–62, doi.org/10.1016/j.jhin.2015.01.017.

10. Aitken M, Gauntlett, C. Patient apps for improved healthcare: from novelty to mainstream. Parsippany (NJ): IMS Institute for Healthcare Informatics; 2013.

11. Price M, Yuen EK, Goetter EM, Herbert JD, Forman EM, Acierno R, et al. mHealth: a mechanism to deliver more accessible, more effective mental health care. Clin Psychol Psychother. 2014;21:427–36, doi.org/10.1002/cpp.1855.

12. Lupton D. Health promotion in the digital era: a critical commentary. Health Promot Int. 2015;30:174–83, doi.org/10.1093/heapro/dau091.

13. Schnall R, Iribarren SJ. Review and analysis of existing mobile phone applications for health care-associated infection prevention. Am J Infect Control. 2015;43:572–6, doi.org/10.1016/j.ajic.2015.01.021.

14. Horsky J, Schiff GD, Johnston D, Mercincavage L, Bell D, Middleton B. Interface design principles for usable decision support: a targeted review of best practices for clinical prescribing interventions. J Biomed Inform. 2012;45:1202–16, doi.org/10.1016/j.jbi.2012.09.002.

15. Van Gemert-Pijnen JE, Nijland N, van Limburg M, Ossebaard HC, Kelders SM, Eysenbach G, et al. A holistic framework to improve the uptake and impact of eHealth technologies. J Med Internet Res. 2011;13:e111, doi.org/10.2196/jmir.1672.

16. Ries E. The Lean Startup: How Today’s Entrepreneurs Use Continuous Innovation to Create Radically Successful Businesses. New York: Crown Publisher; 2011.

17. Willis GB, Artino AR Jr., What Do Our Respondents Think We’re Asking? Using Cognitive Interviewing to Improve Medical Education Surveys. J Grad Med Educ. 2013;5:353–6, doi.org/10.4300/JGME-D-13-00154.1.

18. Stoyanov SR, Hides L, Kavanagh DJ, Zelenko O, Tjondronegoro D, Mani M. Mobile App Rating Scale: A New Tool for Assessing the Quality of Health Mobile Apps. JMIR Mhealth and Uhealth. 2015;3:ARTN e27, doi.org/10.2196/mhealth.3422.

19. Brooke J. SUS-A quick and dirty usability scale. Usability evaluation in industry. 1996;189:4–7.

20. Brooke J. SUS: a retrospective. Journal of usability studies. 2013;8:29–40.

21. Werkgroep Infectie Preventie. Protocol Prevalentieonderzoek Ziekenhuizen [Protocol Prevalance study Hospitals]. 2017

22. Sanger PC, Hartzler A, Han SM, Armstrong CA, Stewart MR, Lordon RJ, et al. Patient perspectives on post-discharge surgical site infections: towards a patient-centered mobile health solution. PLoS One. 2014;9:e114016, doi.org/10.1371/journal.pone.0114016.

23. Fernandes-Taylor S, Gunter RL, Bennett KM, Awoyinka L, Rahman S, Greenberg CC, et al. Feasibility of Implementing a Patient-Centered Postoperative Wound Monitoring Program Using Smartphone Images: A Pilot Protocol. JMIR Research Protocols. 2017;6, doi.org/10.2196/resprot.6819.

24. McGuckin M, Govednik J. Patient empowerment and hand hygiene, 1997-2012. J Hosp Infect. 2013;84:191–9, doi.org/10.1016/j.jhin.2013.01.014.

25. Nielsen J, Landauer TK. A mathematical model of the finding of usability problems. Proceedings of the INTERACT’93 and CHI’93 conference on Human factors in computing systems: ACM; 1993. p. 206–13

